# A rapid review of COVID-19 vaccine prioritization in the U.S.: alignment between Federal guidance and State practice

**DOI:** 10.1101/2021.03.11.21253411

**Authors:** Vageesh Jain, Lara Schwarz, Paula Lorgelly

## Abstract

**Background:** Population groups to be prioritized for COVID-19 vaccinations in the U.S. have been determined at the Federal level. Evidence suggests that there is variation in how States have implemented this guidance. This review examines how the position of population groups in vaccine priority lists varies between Federal guidance and State practice.

**Methods:** An online search of State vaccination prioritization plans was conducted. Data were extracted on each population group included and their relative position. A standardized ranking method was applied to provide a directional measure of variability in prioritization between State and Federal guidance, for each population group.

**Results:** Healthcare workers and those in long-term care facilities were largely prioritized in line with Federal guidance. Aside from early education staff, essential workers were often excluded at State level. Of the 37 States including frontline essential workers, 12 assigned them to a lower priority than recommended. Almost all States included the 65-74 year age group and most assigned them to a higher position than recommended in Federal guidance. Those with underlying medical conditions were similarly highly prioritized, although there was more variability across States. Some socially vulnerable groups (not included in Federal guidance) were highly prioritized by many States.

**Conclusions:** Across the U.S, the prioritization of groups for COVID-19 vaccination has been highly variable. Essential workers were the most often excluded or less highly prioritised group compared to Federal guidance. Some socially vulnerable groups were highly prioritized in State plans, whilst others were mentioned in only a few States. Future guidance must be relevant to local needs and values, to minimise any unwarranted heterogeneity in vaccine access across populations.

## Introduction

Health inequities are differences in health that are unnecessary, avoidable, unfair and unjust^1^. Vertical equity refers to the preferential allocation of resources to those most in need. Although it is expected that all of the United States (U.S.) population will be offered a COVID-19 vaccine eventually, with limited initial supplies, an equitable allocation would involve prioritizing those who are most at need, with particular attention to the socially disadvantaged. Having a robust, scientific and nationally standardized process is essential, as the health and socioeconomic impacts of the pandemic vary greatly across population groups.

Federalism, or the division of power between a national government and States, is a fundamental feature of U.S. public health governance. The Centers for Disease Control and Prevention (CDC) – the leading Federal public health agency in the U.S. – has limited authority to direct local officials to take united action^2^. For example, the power to quarantine individuals rests primarily with State and local authorities, and there is substantial variation between jurisdictions^3^. Despite clear CDC guidance^4^, 13 States still have no mandatory public facemask policy as of March 2021. Arguably, it is possible for a Federalist public health system to respond equitably to a pandemic, but this would require robust Federal guidelines that reflect local needs and values, and their effective implementation.

The population groups to be prioritized for COVID-19 vaccination in the U.S. were determined by the Advisory Committee on Immunization Practices (ACIP), in the CDC. The foremost groups recommended for vaccination ahead of others include healthcare workers and those in long-term care facilities (Table 1). The guidance does not explicitly account for social vulnerability (a risk factor for disease acquisition and severity^5^), instead advising States to consider this at all stages of prioritization. Despite clear Federal guidance, current evidence suggests there is significant variation in the way States are choosing to prioritize population groups for vaccination^6-8^. To some extent this may be warranted given the responsibility of States to tailor central guidance to the needs of their local population. But where there is systematic variability in the position of some population groups, this indicates that Federal guidance is limited in scope and relevance.

**Table 1.** CDC guidance summary and definitions

| Phase | Population Group | Definitions |
| --- | --- | --- |
| **1A** | Healthcare personnel | Paid and unpaid persons serving in healthcare settings who have the potential for direct or indirect exposure to patients or infectious materials |
|  | Long-term care facility residents | Adults who reside in facilities that provide a range of services, including medical and personal care, to persons who are unable to live independently |
| **1B** | Frontline essential workers | First responders (e.g. firefighters and police officers), corrections officers, food and agricultural workers, U.S. Postal Service workers, manufacturing workers, grocery store workers, public transit workers, and those who work in the education sector (teachers and support staff members) as well as child care workers |
|  | People aged 75 and older | People aged 75 and older |
| **1B** | People aged 65-74 years | People aged 65-74 years |
| | People aged 16-64 years with underlying medical conditions | Conditions include cancer; chronic kidney disease; chronic obstructive pulmonary disease (COPD); heart conditions, such as heart failure, coronary artery disease, or cardiomyopathies; immunocompromised state (weakened immune system) from solid organ transplant; obesity (body mass index [BMI] $\geq 30$ kg/m<sup>2</sup> but $< 40$ kg/m<sup>2</sup>); severe obesity (BMI $\geq 40$ kg/m<sup>2</sup>); sickle cell disease; smoking; type 2 diabetes mellitus; and pregnancy |
|  | Other essential workers | Those working in transportation and logistics, water and wastewater, food service, shelter and housing (e.g., construction), finance (e.g., bank tellers), information technology and communications, energy, legal, media, public safety (e.g., engineers), and public health workers |

This paper examines how the position of different population groups in vaccine priority lists varies between Federal guidance and State practice, and which population groups (included in State priority lists) were not included in Federal guidance. This will further our understanding of how Federal guidance on priority groups can be improved, in order to facilitate a more equitable national allocation of vaccines. Given manufacturing and storage limitations, the possibility of novel variants and waning immunity, this study may also inform the rollout of future COVID-19 vaccination programs, as well as future infectious disease vaccination responses.

## Methods

### Definitions and Sources

Recommendations on COVID-19 vaccine priority groups and their definitions, issued at the Federal level by the CDC, were accessed online^9-11^ (Table 1). Currently, this guidance proposes prioritizing seven high-risk population groups in the first phase of vaccination. This phase can be split further into three levels of priority (1A – 1C), with those in 1A receiving vaccines before 1B, and those in 1B receiving vaccines before 1C, although some overlap is expected given variations in populations, group definitions and vaccine supplies. Multiple groups are included within each phase, for instance with both health workers and those in long-term care facilities to be vaccinated in parallel during phase 1A, as the top priority groups. Following the first phase, the remainder of the general population (aged 16+) are to be vaccinated in phase two. No further prioritization within the general population has been so far recommended by the CDC^11^ Definitions of population groups and the phase in which individual States, plus Washington D.C., prioritized them were obtained from publicly available documents, published on official State Department of Health websites.

### Data Extraction

The online search of State guidance was conducted by one reviewer (VJ) from 14-18^th^ February 2021 and repeated by a second reviewer (LS) from 26^th^ February to 2^nd^ March. The second reviewer was blinded to any previous results, but provided with the same data extraction framework used by the first reviewer, to ensure comparability of findings. Where both reviewers recorded the inclusion or position of groups differently, those particular State priority lists were re-visited and it was determined whether guidance had changed between the two independent reviews, or there was a discrepancy in interpretation between reviewers. All such instances were discussed until agreement was reached on the appropriate inclusion or position of groups.

All data were extracted to Microsoft Excel, enabling a comparison of how population groups are prioritized by individual States. All groups specified by individual States were recorded as written in State guidance, including where there were minor differences in the definitions of groups. The position of a particular group in a State priority list was also extracted as written in official guidance, regardless of states numbering groups differently. The resulting database is available from the authors on request.

### Data Analysis

Some groups were aggregated to improve comparability with Federal guidance. This included grouping various age bands (e.g. 65-70 years) into the relevant (75+ or 65-74) groups recommended by the CDC. Those with existing medical conditions (of any age and any number of conditions) were aggregated into one group for analysis due to the use of various age thresholds for those with conditions, across States. Some groups included in State priority lists were explicitly not aggregated for analysis, as considering them distinctly revealed important differences. This included healthcare workers: separated into inpatient and outpatient/community services as well as frontline essential workers, with first responders (in turn stratified into medical and non-medical) and early education staff analyzed separately. In all cases, individual groups were compared with the most relevant Federal guidance benchmark.

The phase in which different groups were positioned by States was converted into a standardized ranking, to negotiate differences in the way States labelled priority groups. For instance some States referred to ‘phase 2’ as the second stage of the vaccine rollout (including the second-most high-priority groups) but in others it was the fourth stage, following three distinct phases from 1A to 1C. For this reason, each population group was considered by whether they were in the first, second, third, fourth or later prioritized phases, for a particular State. This enabled comparison with the three ordered phases set out by Federal guidance (1A-C). Where States included subgroups within a phase (e.g. 1A.1, 1A.2, 1A.3) these were treated as part of the same single phase (1A). Both reviewers involved in data extraction aggregated the data for each population group in this way and discussed any disagreements until consensus was reached.

Overall alignment with Federal guidance was considered in two steps. First, the total number of States including a population group in their vaccination plan was aggregated. Then, using the standardized ranking method outlined above, the total number of States using the same phase as recommended by the CDC, the total number positioning a group ahead of (higher-priority) and the total number positioning a group behind (lower-priority) Federal guidance, was calculated. This provided a directional measure of variability between State practice and Federal guidance, for each included population group.

Many States had explicitly included populations groups that were not covered in the Federal guidance. These groups, as well as the phase of vaccination prioritization plans in which they were included, were summarized separately in a descriptive table.

## Results

Table 2 demonstrates that all U.S States prioritized frontline healthcare workers and those in long-term care facilities for vaccination as per Federal guidance. Unlike other States, Rhode Island included long-term care settings within a wider group initially prioritized for vaccines: congregate settings. Although Federal guidance makes no distinction between healthcare workers based in hospitals or directly caring for COVID-19 patients and others (e.g. pharmacists, dentists), six States assigned a lower priority to those in the latter group. First responders were one single group in Federal guidance, including health (e.g. paramedic) and non-health (e.g. police/fire) workers. In State guidance, these groups were often prioritized differently. Medical first responders were prioritized ahead of the indicated position in Federal guidance, alongside other healthcare workers, in 32 States. Other first responders were more commonly prioritized in phase 1B as per Federal guidance, although still assigned a higher priority position in 14 States.

**Table 2.** State alignment with CDC guidance on COVID-19 vaccine prioritization

| Population group | CDC recommended priority phase | Number of States including in plans (n=51) | Number of States aligned | Number of States assigning higher-priority | Number of States assigning lower-priority |
| --- | --- | --- | --- | --- | --- |
| Frontline health workers | 1A | 51 | 51 | 0 | 0 |
| Long-term care staff/residents | 1A | 50 | 50 | 0 | 0 |
| Other health workers | 1A | 47 | 41 | 0 | 6 |
| First responders (EMS) | 1B | 47 | 12 | 32 | 3 |
| First responders (other) | 1B | 45 | 25 | 16 | 4 |
| Early education staff | 1B | 47 | 38 | 4 | 5 |
| Frontline essential workers | 1B | 37 | 23 | 2 | 12 |
| Age 75+<sup>a</sup> | 1B | 18 | 15 | 2 | 1 |
| Age 65-74<sup>b</sup> | 1C | 47 | 11 | 36 | 0 |
| Age 16-64 with high-risk medical condition<sup>c</sup> | 1C | 40 | 14 | 20 | 6 |
| Other essential workers | 1C | 22 | 15 | 2 | 5 |

<sup>a</sup> includes States where the prioritized age-group was 70+
<sup>b</sup> includes States where the prioritized age-group was 65+ (i.e. no upper limit)
<sup>c</sup> Includes States where high-risk medical conditions group had no age restriction

Essential workers were excluded from many State priority lists. Of the 37 States including frontline essential workers, 12 assigned them to a lower priority than recommended in Federal guidance. Most States included early education staff (including K-12 school and child care workers) at the recommended position, albeit on their own in many cases, rather than as part of the wider essential workforce as recommended by the CDC. Other essential workers (placed in the third phase of Federal recommendations) were included on only 22 State priority lists, although at the recommended Federal position in the majority of cases where present.

Only 18 States included priority age groups of 70 or 75 years and over, with the majority using lower age thresholds. Although 47 States included 65-74 age groups, 36 assigned them to a higher priority than Federal guidance. Those with underlying medical conditions were also commonly included in State priority lists. For this group there was more variability in how they were prioritized, with 20 out of 40 States assigning a higher priority and 6 assigning a lower priority, compared to Federal guidance.

Several other groups were prioritized by States, and not included in Federal guidance (Table 3). Those living or working in congregate settings were explicitly included in the priority lists of 28 States. In the majority of cases they were included in the very first or second phase of a State’s vaccine rollout. Although this group would be expected to include both homeless and prison populations, these terms were explicitly described in only 20 and 9 State priority lists respectively. Individuals living with mental, physical or developmental disabilities were also a commonly identified and highly prioritized group, in 10 States. Other socially vulnerable groups including indigenous or tribal populations, Medicaid beneficiaries, and low-income groups were included in very few State priority lists.

**Table 3.** Other groups prioritized by States

| Population group | Number of States including in plans | Number of States prioritizing first | Number of States prioritizing second | Number of States prioritizing third | Number of States prioritizing fourth or below |
| --- | --- | --- | --- | --- | --- |
| Working/living in congregate settings e.g. prisons, homeless shelters, group homes, emergency shelters, adult/child protective services | 28 | 6 | 13 | 9 | 0 |
| Individuals with disabilities | 10 | 3 | 6 | 0 | 1 |
| Age 50-64 | 7 | 1 | 3 | 1 | 2 |
| Age 60 and over | 5 | 0 | 2 | 3 | 0 |
| Those with moderate-risk conditions | 4 | 0 | 1 | 1 | 2 |
| Indigenous or Tribal populations | 4 | 1 | 1 | 1 | 1 |
| Age 55-64 | 2 | 0 | 0 | 2 | 0 |
| Medicaid beneficiaries | 2 | 0 | 1 | 1 | 0 |
| Low-income/living in affordable housing | 2 | 0 | 2 | 0 | 0 |
| Age 40 and over | 1 | 0 | 1 | 0 | 0 |

## Discussion

### Key findings

For healthcare workers and those in long-term care facilities, there was almost complete concordance between individual State vaccine priority lists and Federal recommendations. Notably first responders were not prioritized in the same way as advised by Federal guidance with medical first responders more often placed ahead of the recommended Federal position and in front of other (non-health) first responders. Many States excluded essential workers from priority lists, and where included they were often assigned a lower priority position compared to Federal guidance. Early education staff was the exception, included in the majority of State priority lists and frequently in the same position as advised by Federal guidance. The majority of States assigned the 65-74 year age group to a higher position than recommended in Federal guidance. Those with underlying medical conditions were similarly highly prioritized, although there was more variability across States. Several socially vulnerable groups not prescriptively included in Federal guidance were often highly prioritized by States, including those living or working in congregate settings and individuals with disabilities.

### Explaining disparities between Federal and State

Several large outbreaks and many associated deaths have been observed in U.S care homes,^12^ emphasising the risk of COVID-19 transmission in such settings. It is therefore unsurprising that staff and residents in long-term care facilities were prioritized by both the CDC and individual States. Healthcare workers are at a high-risk of exposure to COVID-19 and have prolonged contact with vulnerable individuals. Among 2,135,190 people in the UK and USA using the COVID-19 Symptom Study app between March 24 and April 23, 2020, front-line health-care workers had at least a threefold increased risk of reporting a positive COVID-19 test compared with the general community^13^. Front-line healthcare workers who worked in inpatient settings had a much greater risk of infection compared to healthcare workers in other settings. This may in part help to explain why although all States prioritized healthcare workers in the very first phase, a few vaccinated those working in the community after those in hospital.

The elderly and those with co-morbidities are at a high-risk of severe disease and death from COVID-19.^14,15^ Most States, although not all, did prioritize by age to some degree using a range of thresholds. Federal recommendations positioned both the 65-74 year age group and those with underlying medical conditions in the third phase (1C). A majority of State priority lists assigned a higher priority to the former group, and a smaller proportion of States did the same for the latter group. In part this may have been due to a Department of Health and Human Services (DHHS) announcement in January 2021 that encouraged States to vaccinate those over 65 years and all those with high-risk medical conditions.^16^ The magnitude of the risk of severe disease with increasing age is well-established^14^ and likely played a part in the high priority assigned to the 65-74 year age group. Those with underlying conditions were positioned higher than Federal guidance in 20 States. An analysis of over 5 million COVID-19 cases across 3141 U.S counties^17^ found that areas in the most vulnerable quintile (as measured by the CDC Social Vulnerability Index) saw higher rates of COVID-19 cases (rate ratio 2.11, 95% CI 1.97 to 2.26) and deaths (rate ratio 2.42, 95% CI 2.22 to 2.64) compared to the least vulnerable areas. The prevalence of major chronic conditions was 24%–41%11higher in the most vulnerable counties. Prioritizing those with medical conditions may have been an attractive option for States looking to minimize health inequalities, as well as being useful in younger populations where not all high-risk individuals would be identified on the basis of age alone.

Federal guidance states preserving the functioning of society as a goal of the U.S. COVID-19 vaccination program, alongside decreasing death and severe disease as well as reducing the burden of COVID-19 on people already facing disparities ^10^. There was a clear distinction made across States between prioritizing early education staff and other essential worker groups. This is likely due to the high risk of COVID-19 outbreaks in schools, their prolonged closure, and the associated detriment to child learning and development.^18^ States consistently excluded other essential workers from their priority plans. One reason may be due to the size of this group, estimated at around 87 million individuals.^19^ There may also have been overlap with groups previously vaccinated, with the majority of essential workers in the U.S. having at least one underlying high-risk medical condition.^11^ Where included, many States assigned a lower priority for frontline essential workers than advised by Federal guidance. In part this could be due to the relatively low predicted rates of vaccine uptake in the younger working population, relative to other groups^20^ as well as State perceptions about the importance of some vaccination program objectives compared to others.

The COVID-19 case rate has been over four times higher in U.S. prisons compared to the general population.^21^ The impact of COVID-19 in prisons has varied across States. Ohio had a prison mortality rate that was around 11 times the adjusted State whereas six other States reported COVID-19 death rates below adjusted State mortality rates.^21^ This may in part explain why those living in congregate settings were included at a relatively high position in some State’s vaccine rollout plans but not at all in others. Congregate settings also include assisted living facilities, mental health and substance misuse treatment centres, homeless shelters, and others. Individuals in such settings are at a higher risk of disease acquisition^22^ and in many cases severe disease. Vaccinating these socially vulnerable groups during a pandemic can help programs meet principles of equity as well as utility,^23^ likely making them a priority for many despite a lack of prescriptive Federal guidance. While States included some socially vulnerable groups in priority lists, even in the absence of clear Federal guidance, others were excluded. Despite the high burden of pre-existing disease, poverty, systemic and structural discrimination, lack of access to health services and preventative measures, indigenous populations have been relatively ignored in the U.S. pandemic response to date^24^. It appears that this continues to be the case, with only Montana, New Jersey, Oregon and Utah explicitly prioritizing indigenous or tribal populations for COVID-19 vaccination.

### Technology, Cost and Supply

The Federal prioritization helps deliver the CDCs objectives for the COVID-19 vaccination program, including reducing transmission and minimising the risk of morbidity and mortality, minimising the disruption to society and the economy, and ensuring equity in vaccine allocation and distribution. However, these objectives and the resulting prioritization groups may not necessarily reflect the external constraints imposed by the vaccine technology, its supply and the cost of vaccination programs. Subsequent State guidance may reflect these.

Both the Pfizer and Moderna vaccines use mRNA, which needs to be stored at very low temperatures, therefore the logistics for vaccinating with these candidate vaccines are different from the Oxford-AstraZeneca and Johnson & Johnson vaccines, which are easier to deploy and handle. The Pfizer and Moderna vaccines were the first approved vaccines in the U.S. Once this logistic consideration is combined with the need to vaccinate as many people as possible as quickly as possible, then priority groups that can be quickly located (those in care homes, health facilities) may be seen as a more effective and cost-effective target. Arguably those at greatest risk, the socially vulnerable groups, may be harder to identify, target, and uptake may be lower.

It is unclear whether States took the cost of vaccinating into account when operationalizing the Federal priority list to create their own priorities. As part of Operation Warp Speed funding has be made available to States, but this appears to be phased^25^. Likewise the supply constraints of the vaccine may have factored into the States prioritization plans.

### Strengths and Limitations

Unlike previous analyzes of U.S. COVID-19 vaccine priority lists^8^ this rapid review systematically aggregated States across the country by population group. This allowed us to create a directional measure of disparity between Federal guidance and State practice, for each population group, providing us with a better understanding of how Federal guidance is working differently for some groups compared to others, and the potential implications on equity.

A key limitation in our analysis was the inability to measure the time taken to progress through different vaccine allocation phases, and how this might vary across States. Given variable vaccine supplies, group sizes, and ways of labelling priority phases, priority groups may not have been directly comparable. For instance, three States (Connecticut, Florida and Mississippi) prioritized several groups in parallel as part of one single phase. Some other States split their rollout into several, smaller phases. To minimize the impact of this on our analysis, we did not use subgroups within phases where reported (e.g. phase 1A.1, 1A.2, 1A.3). As both Federal and State plans prioritized distinct groups we were able to assess the overall translation of guidance into practice, despite inconsistent methods of prioritization across States. Secondly it was not possible to assess variation in vaccine prioritization at the County level due to a lack of routinely available published documentation. Finally, the extraction and analysis of data across States was limited by the information available in the public domain, with some priority lists being more extensive than others.

### Implications for future research

Disparities between Federal guidance and State practice were inconsistent across population groups. Given the alignment in prioritizing healthcare workers and long-term care facilities, future research is urgently needed to better define social value judgements about vaccine prioritization across the rest of the population. Qualitative research with State decision-makers and citizens, for instance in the form of Citizen Councils^26^, will be necessary to understand how Federal guidance can more accurately reflect local values and objectives. Some States have seen a quicker vaccine rollout compared to others.^6^ This, coupled with the variation in the length of time different groups will wait to receive a vaccine, makes it imperative that the long-term impact on all-cause mortality and morbidity is studied within different population groups. Slower progressing State programs omitting socially vulnerable groups from priority lists (as per Federal guidance) may be considered less equitable, through their failure to protect those who most suffer the socioeconomic impacts of infection or non-immunity, for longer. Given the concentration of ill-health prior to the pandemic, in low-income, deprived and socially disadvantaged groups, this may be worse for health inequalities but has often been justified on the grounds of speed and simplicity.^16^ Nonetheless, other large Federal nations have prioritized socially vulnerable groups differently. For example, Canada prioritizes indigenous communities and marginalized communities in the second phase of their federal vaccine rollout plans. Similarly, Aboriginal adults and those with a disability and specified medical condition are prioritized in the second phase of the federal guidelines of the Australian Department of Health^27^. Understanding the relationship between equity and efficiency in the context of COVID-19 vaccination programs is a key research question that can inform future rollouts, including in countries yet to receive vaccines.

## Conclusions

The prioritization of groups for COVID-19 vaccination has been highly variable across the U.S, despite clear Federal guidance. For healthcare workers and those in long-term care facilities, there is little variation. Other than early education staff, many essential workers were altogether excluded from several State priority plans. Where included they were often assigned a lower priority position compared to Federal guidance. On the other hand, first responders, those aged 65-74 years and those with underlying conditions were frequently assigned to a higher priority position. Socially vulnerable groups such as those living in congregate settings and the disabled were highly prioritized in State plans, despite not forming a distinct priority group in Federal guidance. Few States included indigenous or low-income populations in their lists, suggesting that there is a great deal of variability in social value judgements and perceived needs for vaccines: currently not reflected in Federal guidance. It may also be that the Federal guidance does not reflect the constraints that States are working within, such that the difference in priorities could be due to idealism at the Federal level versus realism at the State level. Future policy must aim to ensure that guidance issued at the Federal level is relevant and sensitive to local needs, acknowledges potential funding and distribution limitations at the local level, and can be implemented in a way that minimises any unwarranted heterogeneity in vaccine access across populations.

## Data Availability

Data available upon request after peer-review and publication

